# Effect of vaccination certification with mass vaccination and non-pharmaceutical interventions on mitigating COVID-19

**DOI:** 10.1101/2023.08.10.23293925

**Authors:** Hu Cao, Longbing Cao

**Affiliations:** School of Computing, Macquarie University, NSW 2109, Australia

## Abstract

As COVID-19 vaccines became abundantly available around the world since the second half of 2021, many countries carried out a vaccination certificate (green pass) policy to encourage vaccination and help reopen their economies. This policy granted certified people more freedom of gathering and movement than unvaccinated individuals. Accordingly, pre-existing non-pharmaceutical interventions (NPIs) were adjusted under the vaccination certificate policy. The vaccination certificate also induced heterogeneous behaviors between unvaccinated and vaccinated groups, which complicates the modeling of COVID-19 transmission. Still, limited work is available in evaluating the impact of the green pass policy on COVID-19 transmission using quantitative methods. To characterize the major changes caused by the green pass policy, a modified susceptible-exposed-infected-removed (SEIR) epidemiological model SEIQRD^2^ is proposed in this paper. By integrating different behavior patterns of unvaccinated and vaccinated groups under the green pass policy, SEIQRD^2^ adopts the inherent variability and complexity of human behaviors in the context of vaccination and NPIs and their effect on COVID-19 transmissions. Three countries: Greece, Austria, and Israel are selected as case studies to demonstrate the validity of SEIQRD^2^. The simulation results illustrate that the combination of NPIs and vaccination still plays a pivotal role in containing the resurgence of COVID-19 by enforcing vaccination certification.

## Introduction

Before December 2020, non-pharmaceutical interventions (NPIs) were the only effective way to contain the spread of the novel coronavirus SARS-CoV-2 [2, 3]. Strict NPIs, including gathering restrictions, border closure, international travel restrictions, curfew, and lockdown, were taken to prevent and contain the COVID-19 pandemic [2]. With the effective vaccines available against SARS-CoV-2 [4], a lot of countries started their nationwide vaccination campaigns since January 2021 [5], including the UK, the US, Israel, and various European countries. Until the second half of 2021, the vaccines were abundantly available around the world. Then, many countries carried out the green pass policy, i.e., individuals and businesses with vaccine passports or certificates have free access to certain facilities and activities. This vaccination certification encouraged people to receive vaccination and helped reopen businesses and economy. For example, Israel was one of the first countries to launch the nationwide vaccination campaign and implement the green pass policy [6].

Since the outbreak of COVID-19, significant efforts have been made on modeling COVID-19 [12], where epidemiological models provide epidemic explanation and indication for controlling the transmission of SARS-CoV-2. In the early stage of the COVID-19 pandemic, the scientific reports focused on the impact of various NPIs on the COVID-19 transmission [7–11]. For example, Jonas et al. [7] discussed the effectiveness of interventions with varying implementation and timing. They showed that strict NPIs and early implementation can largely reduce the spread of COVID-19. Seth et al. [8] made an evaluation of the effect of various NPIs in Europe. Their results demonstrated that lockdowns have a huge impact on reducing COVID-19 transmission. With the development of effective vaccines against SARS-CoV-2, efficient vaccine allocation strategies and the optimal combination of both vaccination and NPIs became valuable topics. Accordingly, a significant number of researchers have developed novel methods or approaches to explore potential combinations [3, 13–23]. For instance, Yang et al. [13] thought that equitable access to vaccine around the world would make a long-term positive impact on curbing COVID-19. Sam et al. [14] investigated different combinations of vaccination and NPIs by varying vaccine’s protection rates against SARS-CoV-2. They concluded that vaccination alone is insufficient to contain the COVID-19 epidemic resurgence. Matrajt et al. [21] searched for potential vaccination strategies to reduce the cumulative cases. Ge et al. [18, 24] evaluated the effectiveness of all kinds of NPIs and vaccination in controlling COVID-19, respectively. Sonabend et al. [23] assessed the roadmap of England by considering the lifting of NPIs and vaccination roll-out under the influence of the COVID-19 pandemic. As the green pass policy has been carried out in several countries, heterogeneous behaviors between vaccinated and unvaccinated groups become another key factor influencing the transmission of COVID-19 pandemic. For instance, the Oxford Covid-19 Government Response Tracker (OxCGRT) distinguished NPIs on vaccinated individuals from the control measures on unvaccinated ones from late July 2022 (https://www.bsg.ox.ac.uk/research/covid-19-government-response-tracker) because of the importation of green pass policy.

NPIs and vaccination affect people’s behaviors significantly, how to model their effect on containing COVID-19 was an emerging topic yet challenging for computing behaviors and their impact [1]. There was some research [39, 40] on considering the impact of heterogeneous behaviors in modeling the dynamics of COVID-19, they are not designed to be applied in the countries or regions where the green pass policy was carried out. For instance, Cliff et al. [39] modeled the development of COVID-19 by incorporating different human behavior patterns under varying environments, such as schools, home, and workplace. Their method makes predictions without carrying out the green pass policy, which cannot differentiate the vaccination status of population and is unsuitable for countries with green pass policy, such as Israel and Greece. A vaccination green pass (vaccination passport or certificate) is a digital or paper certificate that proves its holders being fully vaccinated or recovered after being infected by COVID-19. It grants its holders more freedom of gathering and movement than unvaccinated individuals. In the context of a pandemic resurgence, while unvaccinated individuals are restricted in social activities, those holding green passes are granted entry to various public establishments, including restaurants, bars, cafes, and indoor venues. [25].

Given that the presence of vaccination certification has fostered different behaviors among unvaccinated and vaccinated groups, COVID-19 modeling [12] may benefit from involving behavioral heterogeneity and their modeling their impact on COVID-19 transmissions from behavior informatics perspective [1]. With this motivation, we propose a modified susceptible-exposed-infected-removed (SEIR) epidemiological model SEIQRD^2^. Different from other work that only characterizes the risk of vaccination certificate policy on unvaccinated and vaccinated groups [26, 27], SEIQRD^2^ explicitly characterizes their behavioral heterogeneity and their further interactions simultaneously using two dependent SEIR-based transmission chains. The two transmission chains correspond to the spread of COVID-19 in the unvaccinated and vaccinated, respectively. In the meanwhile, the interplay between the two groups is described by the association between the two chains.

In addition, our model notices the diminishing efficacy of vaccination during the COVID-19 pandemic. Differently, many research studies on COVID-19 do not consider the decreasing effect of vaccination on infection [18, 19, 28]. This ignorance may conflict with the reality of the COVID-19 resurgence, to which the waning effectiveness of vaccines made a contribution [29–32].

This work makes the attempt to precisely explore the above open question per the following real-world scenario and assumptions. First, authorities would relax NPIs and grant the vaccinated group more freedom to activities such as shopping and public gatherings than the unvaccinated [25]. Second, the virus resurgence could be triggered after the implementation of the green pass policy. These assumptions represent the practical situations in the countries which implemented the green pass policy in 2021. Still, limited work is available in evaluating the impact of the green pass policy on people’s behaviors and the COVID-19 transmission from a quantitative perspective. This paper addresses these gaps in SEIQRD^2^.

SEIQRD^2^ analyzes the above aspects and gaps in modeling the viral and human behavior dynamics during COVID-19 under mass vaccination and the influence of vaccination certification and other NPIs. SEIQRD^2^ also captures the behavioral heterogeneity between vaccinated and unvaccinated groups under the green pass policy characterized by two dependent transmission chains. At last, we demonstrate the validity of this method in modeling COVID-19 transmissions in three pioneering countries with vaccination certification: Greece, Austria, and Israel.

## Materials and methods

### Model structure and assumptions

The structure of SEIQRD^2^ is shown in Fig 1, which illustrates the organization and arrangement of various components of the model. The left part of the figure displays major external factors for COVID-19: virus, mass vaccination, and NPIs. The right section outlines the dynamics of COVID-19. As it is shown, the compartmental transition part explicitly incorporates the impact of these external factors. For instance, the virus SARS-CoV-2 caused the COVID-19 pandemic and its transmission could be described by an extended SEIR model. In a mass vaccination campaign, the total population is typically categorized into two groups: those who have been vaccinated and those who have not received the vaccine (unvaccinated). Lastly, when a green pass policy was carried out, stricter NPIs were used to restrict the activities of the unvaccinated group than the vaccinated, which induced heterogeneous behaviors between the two groups. All these factors form the two dependent SEIR-based chains in SEIQRD^2^.

**Fig 1.**
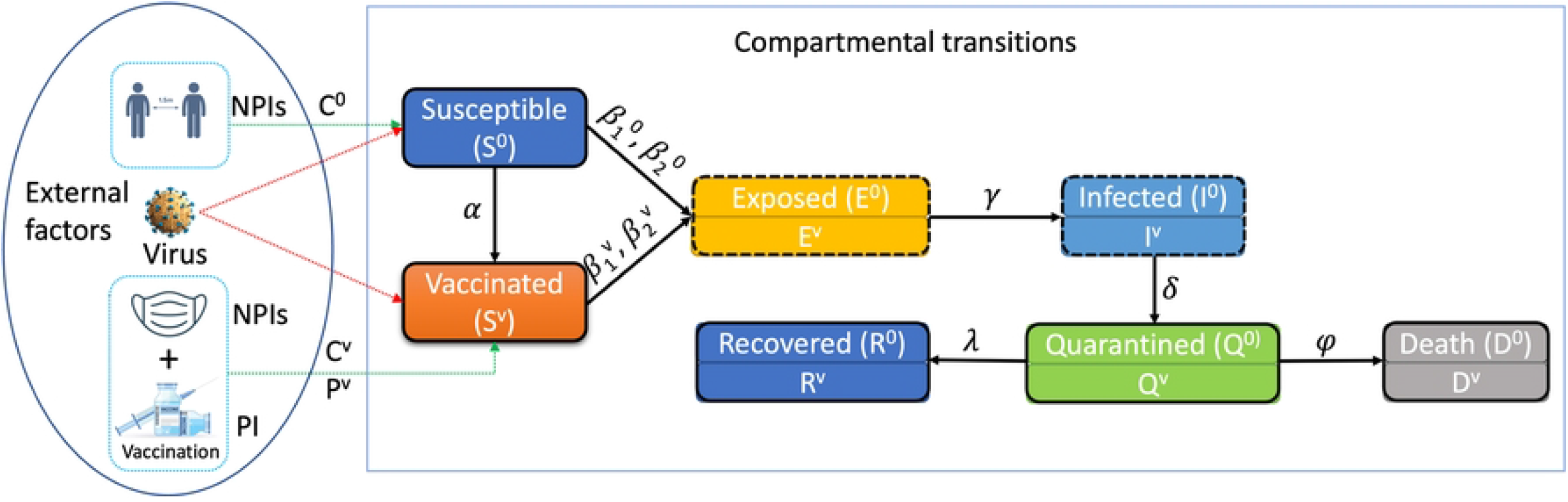
The structure and workflow of the model SEIQRD^2^. In the external factors part, C^0^ and C^*v*^ represent NPI events for the unvaccinated and vaccinated, respectively; P^*v*^ stands for mass vaccination campaign. In the compartmental transitions part, the status transitions among unvaccinated population are described by the chain S^0^, E^0^, I^0^, Q^0^, R^0^, and D^0^. Similarly, the transition chain of the vaccinated consists of S^*v*^, E^*v*^, I^*v*^, Q^*v*^, R^*v*^, and D^*v*^. *α* is the daily vaccination rate, 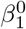 represents the effective transmission rate among Susceptible(S^0^) caused by Infected (I^0^), 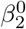 depicts the effective transmission rate among Susceptible(S^0^) caused by Infected (I^*v*^), 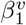 is the effective transmission rate among Vaccinated(S^*v*^) caused by Infected (I^0^), 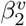 is the effective transmission rate among Vaccinated(S^*v*^) caused by Infected (I^*v*^), 1/*γ* is the incubation period, *δ* is the diagnosis rate, *λ* is the recovery rate, and *φ* is the death rate.

### Compartmental transitions

The compartmental transitions part comprises two similar transition chains. The first chain is for the unvaccinated people. It consists of seven compartments: Susceptible (S^0^), Exposed (E^0^), Infected (I^0^), Quarantined (Q^0^), Recovered (R^0^), and Death (D^0^), which reflect the epidemiological compartmental transitions among the unvaccinated individuals during COVID-19. These components are presented in Fig 1. Individuals in the Susceptible compartment (S^0^) may get infected by those Infected (I^0^) or Infected (I^*v*^) at the effective transmission rate of 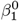 and 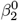, and become exposed, i.e., transited to the Exposed status (E^0^). After a particular incubation period 1/*γ*, those exposed individuals (E^0^) are transferred to the Infected compartment (I^0^) and become infectious. Those infectious may be identified and confirmed, then transferred to the Quarantined compartment (Q^0^) at a probability *δ*. In Q^0^, individuals either recovers (R^0^) at rate *λ* or deceases (D^0^) at rate *φ*.

The other transition chain is for those vaccinated with green passes. Similarly, it consists of compartments Vaccinated (S^*v*^), Exposed (E^*v*^), Infected (I^*v*^), Quarantined (Q^*v*^), Recovered (R^*v*^), and Death (D^*v*^). The set of compartments depicts the dynamics of COVID-19 among green pass holders. Besides, the transition processes are also similar to those unvaccinated people (i.e., the status S^0^, E^0^, I^0^, Q^0^, R^0^, and D^0^). The compartments S^*v*^, E^*v*^, I^*v*^, Q^*v*^, R^*v*^, and D^*v*^ are shown in Fig 1.

This bi-chain compartmental structure describes the status transitions among unvaccinated and vaccinated people during the COVID-19 pandemic and grasps their behavioral heterogeneity simultaneously. Different from the existing epidemiological methods [13, 14], SEIQRD^2^ emphasizes the interactions between the two groups, which play an important role in forming the resurgence of COVID-19 under the green pass policy. For instance, a susceptible person (S^0^) may become infected by those vaccinated (I^*v*^) at the probability of 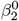. Similarly, 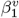 reflects the transmission rate that the unvaccinated (I^0^) spreads the virus to a green pass holder (S^*v*^). The two dependent SEIR-based chains of SEIQRD^2^ can be characterized by the following set of ordinary differential equations (ODE) Eqs (1)-(12). *N* refers to the population size.

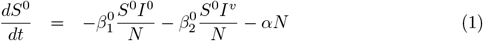

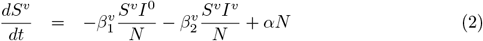

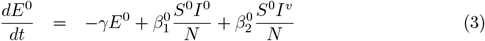

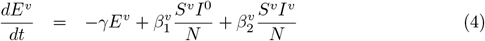

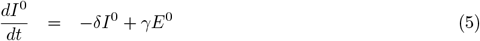

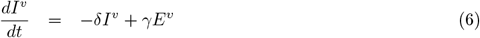

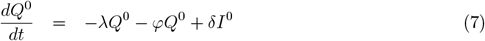

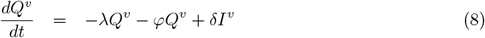

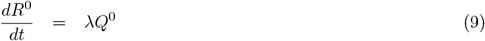

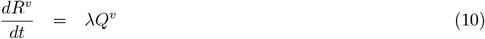

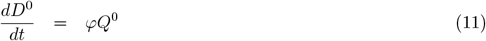

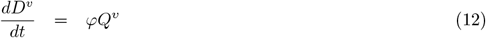

*α, λ, φ* are country specific parameters from real-world data described in Table 1, while 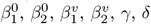 are estimated from the model.

**Table 1.**
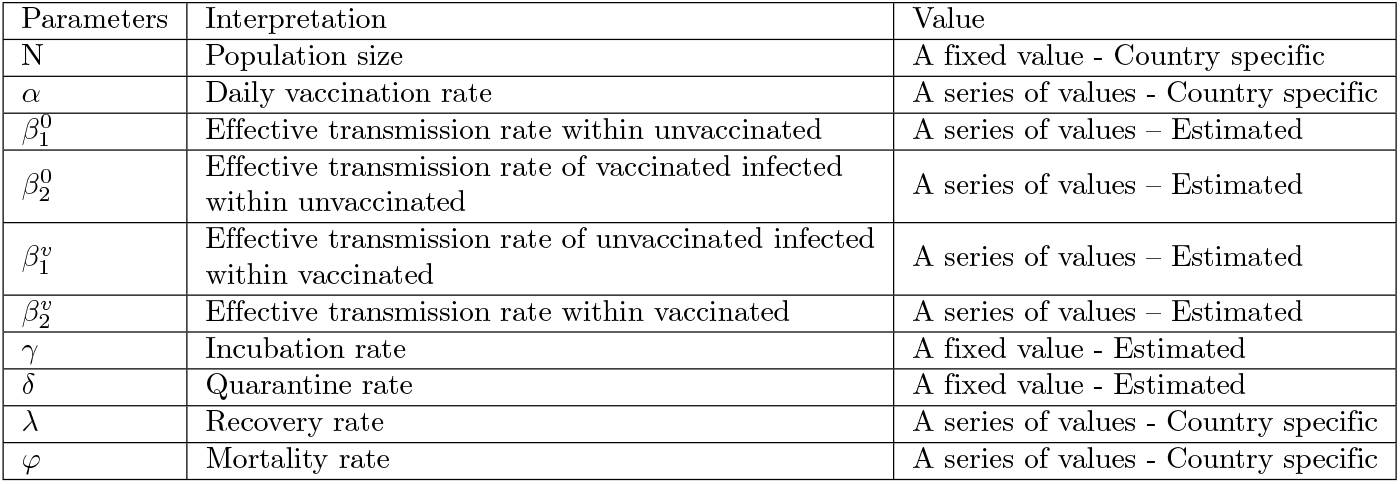
Model parameters for SEIQRD^2^ and their associated values.

| Parameters | Interpretation | Value |
| --- | --- | --- |
| N | Population size | A fixed value - Country specific |
| $\alpha$ | Daily vaccination rate | A series of values - Country specific |
| $\beta_1^0$ | Effective transmission rate within unvaccinated | A series of values – Estimated |
| $\beta_2^0$ | Effective transmission rate of vaccinated infected within unvaccinated | A series of values – Estimated |
| $\beta_1^v$ | Effective transmission rate of unvaccinated infected within vaccinated | A series of values – Estimated |
| $\beta_2^v$ | Effective transmission rate within vaccinated | A series of values – Estimated |
| $\gamma$ | Incubation rate | A fixed value - Estimated |
| $\delta$ | Quarantine rate | A fixed value - Estimated |
| $\lambda$ | Recovery rate | A series of values - Country specific |
| $\varphi$ | Mortality rate | A series of values - Country specific |

### External factors

Compartmental transitions may be greatly affected by external factors: NPIs and mass vaccination. Therefore, it is necessary for SEIQRD^2^ to quantify the effect of these NPI events and vaccination. Generally, the impact of such external factors is assessed by their role in changing the viral transmission speed. In SEIQRD^2^, we define the effect of external factors NPIs and vaccination by measuring their impact on transmission rates. The effective transmission rates 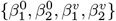 are described by Eq (13). Each transmission rate is a function of NPIs and vaccination.

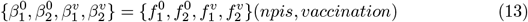

In theory, NPIs and vaccination are independent from each other. Therefore, Eq (13) is transformed into Eq (14).

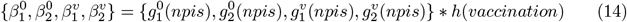

We further explain the functions of 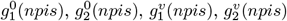, and *h*(*vaccination*). During the COVID-19 pandemic, various NPIs were carried out to stop or mitigate the viral spread. Commonly, step functions are used to measure the effect of existing NPIs during a COVID-19 resurgence. In the other words, at a given time segment, the effectiveness of NPIs is fixed as a constant value. The discrete values come from fitting the model, where *n* is the total number of NPIs-related events. *T*_*i*_ represents the starting time of the i^*th*^ NPI event, *T*_*total*_ is the total number of days during a specific COVID-19 wave. For example, 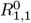 represents the impact of NPIs among the unvaccinated after the 1^*th*^ NPI event and before the 2^*nd*^ event, called basic transmission rate. Therefore, the functions of NPIs can be characterized as a set of discrete values by Eqs (15)-(18).

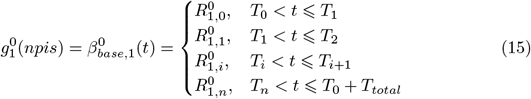

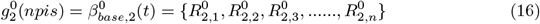

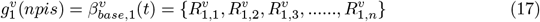

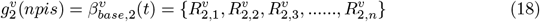

The effectiveness of vaccination is measured by Eq (19). Many scientific studies explore the effectiveness of vaccines against the infection of COVID-19 over time. In this work, we use the results about the Pfizer–BioNTech (BNT162b2) COVID-19 vaccine in [33] to compute the effectiveness of vaccination during COVID-19 resurgence because the Pfizer vaccination took the highest proportion around the world, including the chosen three countries: Greece (*>* 70%), Austria (*>* 60%), and Israel (*>* 80%). *M* refers to the total population of the vaccinated until the t^*th*^ day. *t*_0_ is the starting time of mass vaccination. *Num*(*j*) is the number of the vaccinated people on the *j*^*th*^ day. *Eff* (*t − j*) refers to the vaccine effectiveness against infection (*t − j*) days after vaccination. In Eq (20), the coefficients {0.775, 0.732, 0.696, 0.517, 0.225, 0.173} correspond to the results in [33]. To simplify the modeling, we assume the booster’s effectiveness follows the same diminishing efficacy trend in [33]. Besides, the assumption will not significantly change the general results of SEIQRD^2^ about NPIs, because the impact of NPIs is described by step functions Eqs (15)-(18), while the effectiveness of vaccination is represented by the ramp function Eq (19), which means it is easy to separate the impact of NPIs from vaccination.

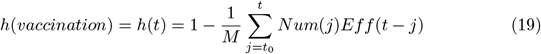

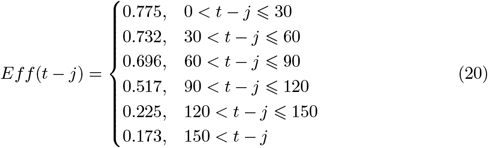

Then, Eq (14) can be converted to Eqs (21-24).

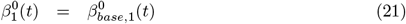

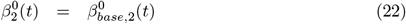

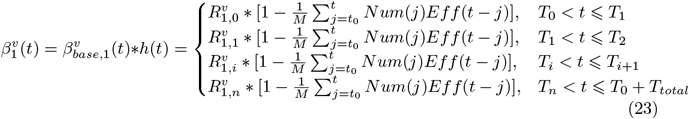

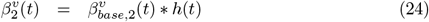

### Estimation

We solve the model with a nonlinear data-fitting approach that minimizes a least squares error function as shown in Eq (25), where *F* represents our model, **x** denotes the input data (*Q, R* and *D*), provided by the integration of the ordinary differential equation (ODE) system (Eqs (1)-(12)) and solved by a fourth-order Runge-Kutta method. *y* denotes an observation (i.e., the reported active case number). *θ* refers to all parameters 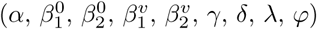 that are inferred by Eqs (1)-(12). *T*_*total*_ is the total number of days.

Model parameters are based on formal estimation. In order to obtain the simulation results, we take the classical optimization algorithm (the least-square method) to fit actual quarantined cases with a given objective function Eq (25). In Eq (25), *y*_*j*_ is the actual quarantined case in the j^*th*^ day.

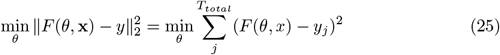

This function requires initial values for optimization. We randomly set the initial values for the unknown parameter set *θ* with the lower bound 0. The representative measure of the optimal set of parameters is obtained with up to 10,000 iterations of optimization under the initial values.

### Evaluation

The MAPE defined in Eq (26) is also a key metric to assess the performance of SEIQRD^2^ in predicting the dynamics of COVID-19. *y*_*j*_ is the actual quarantined case or total deaths on the j^*th*^ day. 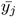 is the prediction of SEIQRD^2^ about quarantined cases or total deaths. *T*_*total*_ is the total number of days. For instance, the prediction of the next 14-day quarantined cases and total deaths is measured by the MAPE of the simulation results.

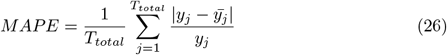

### Explanatory model for transmission rates

As we know, classic SEIR models are widely applied in the public health domain. One of the most obvious reasons is its clear explanation. It means they can provide key epidemiological parameters, such as transmission rate and incubation period. In our model, four effective transmission rates 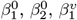, and 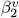 depict a more accurate dynamics of the pandemic by specifying people’s vaccination status. Unlike the single transmission rate in classic SEIR models, the four effective transmission rates equip the model with a more accurate picture about the COVID-19 transmission. For example, the increase of both 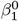 and 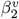 reflects the relaxation of NPIs or the appearance of a more infectious virus variant. Combined with the decrease of 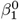, the increase of 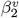 points out that the phenomenon may be caused by over-relaxation interventions for green pass holders or declined vaccine effectiveness. Furthermore, 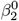 and 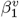 capture the interactions between unvaccinated and vaccinated groups. For instance, the rise of 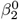 shows that the activities of those vaccinated increase the risk of unvaccinated people being infected. Accordingly, it would be reasonable to enforce appropriate restrictions on those vaccinated. The change of 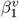 may be caused by lifting NPIs for the vaccinated and the waning efficacy of vaccination. With these four rates, SEIQRD^2^ can offer a more comprehensive understanding about the dynamics of COVID-19 both within and between the groups, which is helpful to adjust the existing control measure reasonably.

The effective transmission rates measure the joint impact of NPIs and mass vaccination on the COVID-19 transmission, while its corresponding basic transmission rates 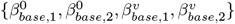 are separated from mass vaccination and consider the impact of NPIs alone. The basic transmission rates are important to evaluate the effectiveness of NPIs when mass vaccination is available.

## Results

### Data and code availability

Data used in the paper are collected from several public datasets. For example, epidemiological data are from the Johns Hopkins University Center for Systems Science and Engineering (JHU CSSE) [34, 35], including daily case numbers and deaths. The vaccination information comes from Our World in Data [36]. The Oxford COVID-19 Government Response Tracker (OxCGRT) [37] provides NPI information and CoVariants [38] with viral variants.

Besides, the data for the second half of 2021 are selected for two reasons. Firstly, a growing number of countries started to carry out green pass policy several months after mass vaccination campaigns started, which happened in this period. Secondly, most countries relaxed COVID-19 monitoring or did not record daily cases since the Omicron variant emerged and became dominant in the late December 2021, which led to inaccuracy of confirmed cases after this period. Based on the above reasons, the data from July to December 2021 are suitable for our research purpose. The data and code are available at GitHub (https://github.com/lzxiaohu/SEIQRD2).

We apply SEIQRD^2^ in three countries: Greece, Austria, and Israel to evaluate its performance. These three countries distributed in Europe and Middle East hold different ethnic and cultural backgrounds, which promote their varied behaviors and responses to NPIs, vaccination, and green pass policy. It could be used to test the generalized ability of our model in different countries. On the other hand, there are several similarities among Greece, Austria, and Israel. They are developed countries and own a sufficient amount of vaccines for their nationwide mass vaccination. They all carried out the green pass policy to restart economic activities when a high percentage of vaccination was reached, which matches with the assumptions of the model. Below, we make a summary of the main results and findings of the experiments.

### Greece

The results for Greece are shown in Fig 2. From 18 September 2021, Greece experienced a new resurgence caused by the Delta variant after a mass vaccination campaign. This wave lasted until late December 2021. Fig 2. A displays the forecasting results about quarantined cases and total deaths for the next 14 days. The accuracy of prediction is measured by the metric MAPE, achieving 3.229% in quarantined cases and 4.853% in death cases. Fig 2B shows the combined impact of NPIs and mass vaccination during the COVID-19 resurgence.

**Fig 2.**
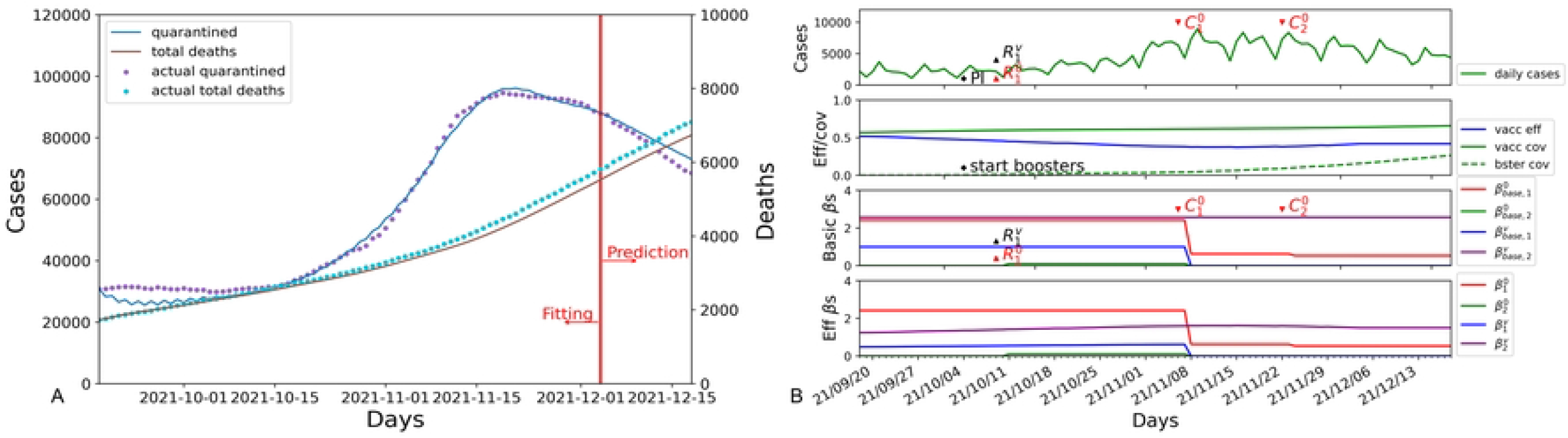
The results of SEIQRD^2^ in Greece for the period between 18 September 2021 and 18 December 2021. (A) is the modeling results for the quarantined and total deaths in Greece, where the blue line refers to the quarantined cases estimated by SEIQRD^2^, and the brown line is the total death cases produced by our method. (B) displays the impact of external factors during the COVID-19 resurgence. The first subplot from the top refers to the daily reported cases in Greece. Vaccination coverage, booster coverage, and the efficacy of vaccination are shown in the second subplot. The third subplot indicates the impact of NPIs on the transmission of the COVID-19 wave, in which the red line is the basic transmission rate 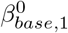 in the unvaccinated group, the green line refers to the basic transmission rate 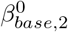 among the unvaccinated caused by the vaccinated, the blue line represents the basic transmission rate 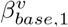 among the vaccinated group caused by the unvaccinated people, and the purple line shows the basic transmission rate 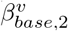 of the vaccinated. The fourth subplot shows the joint impact of both NPIs and vaccination during the COVID-19 resurgence, which are denoted by the four effective transmission rates 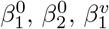, and 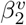. PI represents the start of booster campaign. 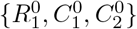 denote the NPI events for the unvaccinated, where *R* means relaxation of NPI and *C* is the control of NPI. Similarly, 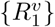 is the NPI event for the vaccinated. Their physical meanings are also listed in the appendix.

As shown Fig 2B, the turning point of the COVID-19 wave appeared on about 09 November 2021, which coincided with the evident decline of effective transmission rates 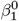 and 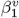 from 2.418 and 0.482 to 0.625 and almost 0, respectively. Combined with the NPI event 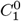 that required unvaccinated individuals to show a negative COVID-19 test result before entering all indoor public places, the wave was contained by the stricter control measures on the unvaccinated group, which slowed down the transmission speed among the unvaccinated and indirectly mitigated the spread among the green pass holders by significantly reducing the physical contact between the two groups. Besides, the decrease of 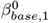 and 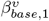 further demonstrates the NPI event 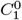 largely reduced the spread of COVID-19. After this, another NPI event 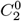 that banned unvaccinated individuals from accessing restaurants came into effect on 22 November 2021. The new NPI further restricted the social activities of those unvaccinated, leading to the decline of transmission rate 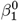. Combined 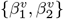 with 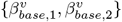, these NPI events did not obviously change the dynamics of the COVID-19 among the green pass holders due to green pass policy. In order to explain the impact of NPI events during the resurgence in detail, we make a table to list their contribution to the four couples of transmission rates. The content is shown in the table 2.

**Table 2.** Attributes of NPI impact in Greece.

| NPI events | Basic transmission rates |  |  |  | Effective transmission rates |  |  |  |
| --- | --- | --- | --- | --- | --- | --- | --- | --- |
| | $\beta_{base,1}^0$ | $\beta_{base,2}^0$ | $\beta_{base,1}^v$ | $\beta_{base,2}^v$ | $\beta_1^0$ | $\beta_2^0$ | $\beta_1^v$ | $\beta_2^v$ |
| Initial values | 2.418 | 0.001 | 1.000 | 2.577 | 2.418 | 0.001 | 0.482 | 1.164 |
| $R_1^0$: 09 Oct 2021;<br>$R_1^v$ | 2.418 | 0.102 | 1.000 | 2.577 | 2.418 | 0.102 | 0.541 | 1.394 |
| $C_1^0$: 06 Nov 2021<br>-: | 0.625 | 0.001 | 0.001 | 2.577 | 0.625 | 0.001 | 0.001 | 1.592 |
| $C_2^0$: 22 Nov 2021<br>-: | 0.531 | 0.001 | 0.001 | 2.577 | 0.531 | 0.001 | 0.001 | 1.599 |

During the resurgence, the government of Greece started a mass booster campaign to mitigate the transmission of COVID-19. This scenario is reflected in the efficacy curve of vaccination shown in Fig 2B. Because of the waning efficacy of vaccination [33], the efficacy of vaccination was decreasing from 0.519 (18 September 2021) to 0.373 (15 November 2021), which caused the increase of the effective transmission rate 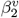. As the acceleration of booster shots happened on 15 November 2021, its effectiveness gradually increased and reached about 0.500 (06 December 2021), corresponding to 1.289 (2.577*0.5) of 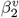. The increase of its efficacy contributed to the mitigation of COVID-19 among the green pass holders, which coincided with a drop in daily cases.

According to the results of SEIQRD^2^, the mitigation of COVID-19 wave was attributed to both control measures and booster shots. In Greece, NPIs were used to reduce the COVID-19 transmission in the unvaccinated group, whereas vaccine booster was for the vaccinated individuals.

### Austria

The modeling results for Austria are shown in Fig 3. A resurgence with more infected cases took place in the middle of September 2021. SEIQRD^2^ makes predictions about the dynamics of the COVID-19 wave for the next 14 days, showing 5.411% MAPE of forecasting quarantined cases. They are illustrated in Fig 3A. The joint impact of external factors is displayed in Fig 3B.

**Fig 3.**
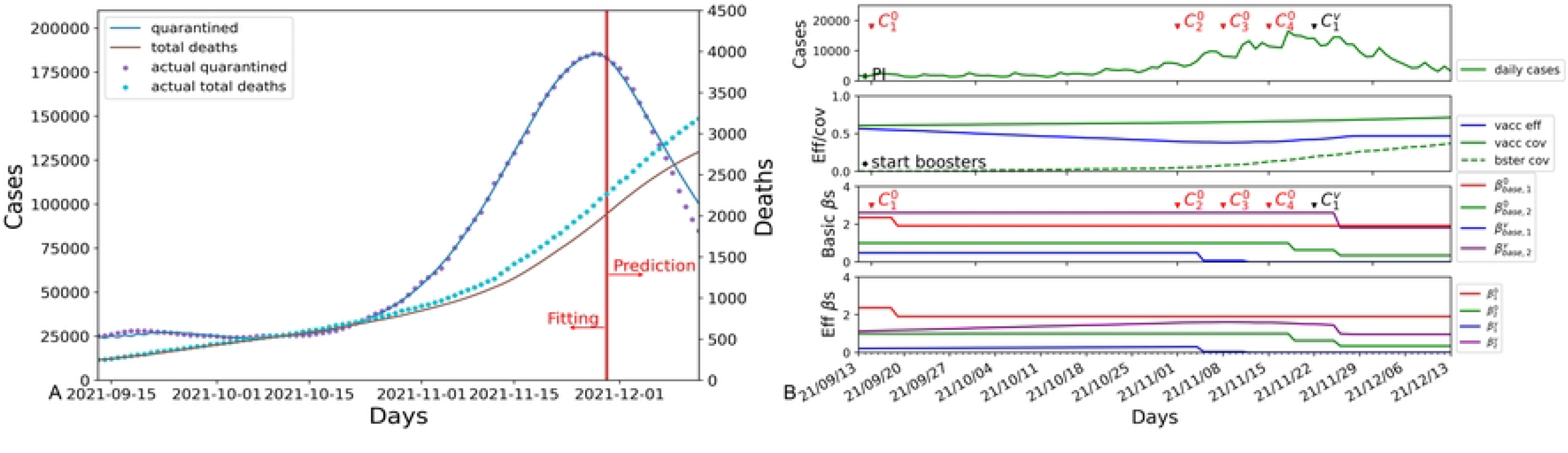
The results of SEIQRD^2^ in Austria for the period between 13 September 2021 and 13 December 2021. (A) is the modeling results for the quarantined and total deaths in Austria. (B) displays the impact of external factors during the COVID-19 resurgence.

The dynamics of the wave is fully explained by SEIQRD^2^ as shown in Fig 3B. After the peak daily cases reported on 18 November 2021, the wave reached a plateau and then declined sharply on 29 November 2021. It was closely related to two lockdowns taken place on 15 November 2021 and 22 November 2021, respectively. In the first lockdown, the government of Austria required all unvaccinated individuals to stay at home 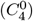, which nevertheless did not quickly reduce the number of infection cases but reduced the physical contact between the two groups. It lowered the transmission rate 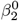 from 0.999 to 0.632. Then, their authorities carried out the second lockdown 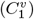 to restrict the social activities of those green pass holders. This second lockdown reduced the values of 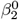 and 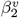 from 0.632 and 1.588 to 0.345 and 1.048, corresponding to 0.345 of 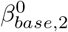 and 1.817 of 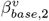. Compared to the first lockdown enforced on the unvaccinated group, the second one on green pass holders made a greater contribution to flattening the wave. That implied that the resurgence may be caused by the over-relaxation of NPIs, especially for those green pass holders. The impact of other NPI events is listed in Table 3.

**Table 3.** Attributes of NPI impact in Austria.

| NPI events | Basic transmission rates |  |  |  | Effective transmission rates |  |  |  |
| --- | --- | --- | --- | --- | --- | --- | --- | --- |
| | $\beta_{base,1}^0$ | $\beta_{base,2}^0$ | $\beta_{base,1}^v$ | $\beta_{base,2}^v$ | $\beta_1^0$ | $\beta_2^0$ | $\beta_1^v$ | $\beta_2^v$ |
| Initial values | 2.365 | 1.000 | 0.487 | 2.607 | 2.365 | 1.000 | 0.211 | 1.130 |
| $C_1^0$: 15 Sep. 2021<br>-: | 1.915 | 0.999 | 0.487 | 2.607 | 1.915 | 0.999 | 0.214 | 1.145 |
| $C_2^0$: 01 Nov. 2021<br>-: | 1.915 | 0.999 | 0.070 | 2.607 | 1.915 | 0.999 | 0.042 | 1.577 |
| $C_3^0$: 08 Nov. 2021<br>-: | 1.915 | 0.999 | 0.001 | 2.607 | 1.915 | 0.999 | 0.001 | 1.610 |
| $C_4^0$: 15 Nov. 2021<br>-: | 1.915 | 0.632 | 0.001 | 2.607 | 1.915 | 0.632 | 0.001 | 1.588 |
| -:<br>$C_1^v$: 22 Nov. 2021 | 1.915 | 0.345 | 0.001 | 1.817 | 1.915 | 0.345 | 0.001 | 1.048 |

Besides, vaccination also played a crucial role in the mitigation of COVID-19. As the resurgence worsened, booster shots received greater attention in public discourse and booster coverage grew substantially from November 2021. Until 13 December 2021, it indeed improved the effectiveness of vaccination from the lowest point of 0.382 to 0.551. After the second lockdown, the effective transmission rate 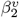 gradually decreased from 1.048 to 0.816 (1.817*(1-0.551)) because of the increasing effectiveness of vaccination.

The results of SEIQRD^2^ demonstrated that proper NPIs are still necessary for the vaccinated group, especially when the efficacy of vaccination decreases. At the same time, vaccination is also critical to control the spread of COVID-19.

### Israel

Lastly, Fig 4 illustrates the modeling results for Israel. Israel was hit by another wave from August 2021 to October 2021. The results modeled by SEIQRD^2^ match the dynamics of this viral resurgence. The MAPE of forecasting the quarantined cases is 1.292% and the MAPE for estimating the total deaths is 3.167%. Fig 4B shows the combined impact of NPIs and mass vaccination during the COVID-19 resurgence in Israel.

**Fig 4.**
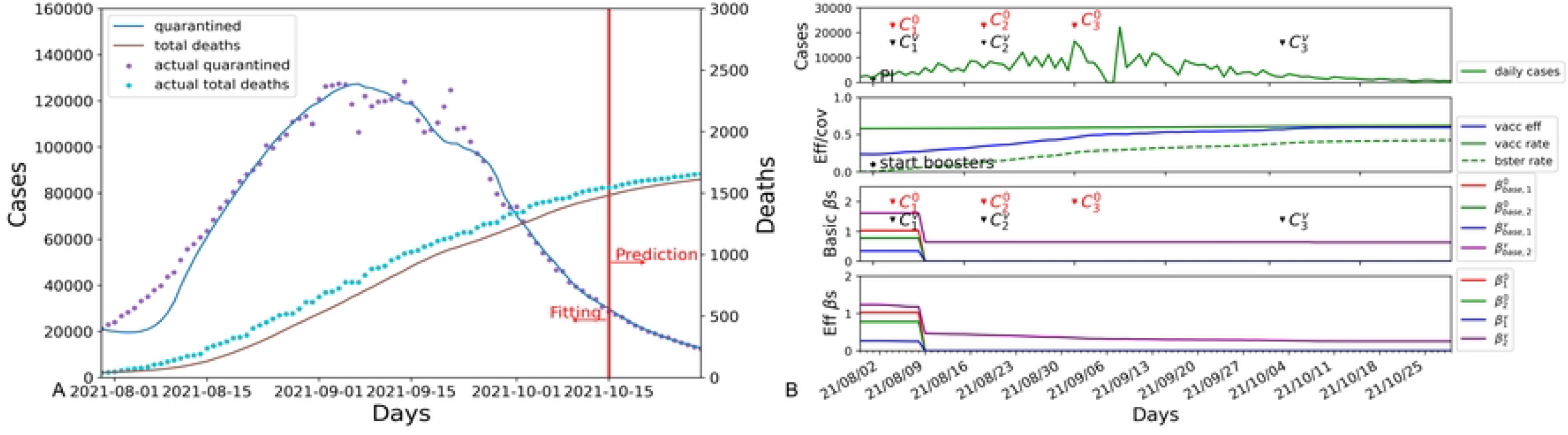
The results of SEIQRD^2^ in Israel for the period between 30 July 2021 and 30 October 2021. (A) is the modeling results for the quarantined and total deaths in Israel. (B) is the impact of external factors on the COVID-19 resurgence.

Then, Fig 4B about the transmission rates of COVID-19 elucidates the course of the resurgence. At the outbreak of this new wave, the Israeli government took stringent NPI measures to control the spread of viral infections. These control measures include showing a negative COVID-19 test result for those unvaccinated 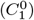 and limiting the gatherings of green pass holders 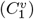, which substantially reduced the four basic transmission rates 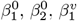, and 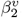. For example, 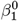 and 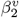 decreased radically from 1.024 and 1.236 to 0.001 and 0.486, respectively. It means that only green pass holders can take part in social activities with limitations, whereas the unvaccinated people were required to stay at home, which described the realistic condition in Israel during the viral resurgence. As a result, the physical contact between the two groups declined substantially, which is demonstrated by the reduction of 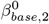 and 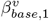 from 0.779 and 0.350 to 0.001 and 0.001. The impact of NPI events is shown in Table 4.

**Table 4.** Attributes of NPI impact in Israel.

| NPI events | Basic transmission rates |  |  |  | Effective transmission rates |  |  |  |
| --- | --- | --- | --- | --- | --- | --- | --- | --- |
| | $\beta_{base,1}^0$ | $\beta_{base,2}^0$ | $\beta_{base,1}^v$ | $\beta_{base,2}^v$ | $\beta_1^0$ | $\beta_2^0$ | $\beta_1^v$ | $\beta_2^v$ |
| Initial values | 1.024 | 0.779 | 0.350 | 1.621 | 1.024 | 0.779 | 0.112 | 1.236 |
| $C_1^0$: 04 Aug. 2021<br>$C_1^v$: | 0.001 | 0.001 | 0.001 | 0.648 | 0.001 | 0.001 | 0.001 | 0.486 |
| $C_2^0$: 18 Aug. 2021<br>$C_2^v$: | 0.001 | 0.001 | 0.001 | 0.647 | 0.001 | 0.001 | 0.001 | 0.425 |
| $C_3^0$: 01 Sep. 2021<br>-: | 0.001 | 0.001 | 0.001 | 0.647 | 0.001 | 0.001 | 0.001 | 0.348 |
| -:<br>$C_3^v$: 03 Oct. 2021 | 0.001 | 0.001 | 0.001 | 0.637 | 0.001 | 0.001 | 0.001 | 0.274 |

In the meantime, Israel started its mass booster campaign to strengthen the effectiveness of vaccination. The turning point of daily cases appeared when the efficacy of vaccination was rising from 0.236 to 0.596. Until the end of COVID-19 resurgence, the value of 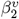 was 0.257 (0.637*(1-0.596)). Therefore, accelerating boosters in a short time period played an important role in suppressing the spread of this COVID-19 wave. Consequently, Israel successfully contained the COVID-19 resurgence through the combination of efficient NPIs and mass booster vaccination within three months.

### Analyses

During the viral resurgences caused by the Delta variant, all three countries: Greece, Austria, and Israel took similar actions to combat COVID-19. These measures comprise a range of different operations, including accelerating booster shots, green pass policy, and more stringent NPIs. Displayed in the results of the model SEIQRD^2^ in three countries, booster shots and green pass policy are also carried out. Furthermore, booster vaccination and stricter NPIs indeed played a crucial role in preventing the transmission of COVID-19. For example, Israel was most successful among these countries in stopping the spread of COVID-19. In the early stage of resurgence, the Israeli government carried out a series of strict NPIs 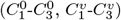 for both the unvaccinated and green pass holders, which largely lowered the transmission of COVID-19. After that, a nationwide booster campaign was activated and the booster coverage increased quickly from 0.5% to 24.5% within a month, improving vaccination efficacy from 0.236 to 0.45. The other two countries adopted a similar combination of NPIs and booster campaign, while their NPI policies were mild and their processes of booster shots were delayed.

On the other hand, the impact of vaccination and NPIs play a crucial role in the development of COVID-19 resurgence in different mechanisms. The effectiveness of vaccination is determined by vaccination rate and vaccine effectiveness against infection. It could only provide protection for the vaccinated. While the influence of NPIs can be varying and unpredictable. According to the simulation results above, even the lockdown were carried out in Greece, Austria, and Israel, their impact on the transmission rates is different. For example, the lockdown 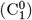 almost stopped the transmission of COVID-19 in the unvaccinated group of Israel, while it 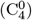 made no difference in Austria. Besides, the NPIs for a specific group, such as the unvaccinated and the vaccinated, would influence the COVID-19 transmission in the other one. For example, the NPI event 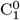 of Greece and 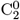 of Austria also change the transmission of COVID-19 within the vaccinated through reducing the effective transmission rate 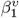 from 0.541 to 0.001, 0.214 to 0.042, respectively. That implies that the impact of NPIs on COVID-19 are also complicated when green pass policy is in effect, which could be captured in the model SEIQRD^2^. To further illustrate the impact of green pass policy, we list the total number of infected and deaths when the unvaccinated people shared the same freedom as the vaccinated and compare it to actual confirmed cases and deceases when green pass policy was available. The detailed information is shown in Table 5. The results suggest that green pass policy reduces the total confirmed cases and total deaths more than 10 times.

**Table 5.**
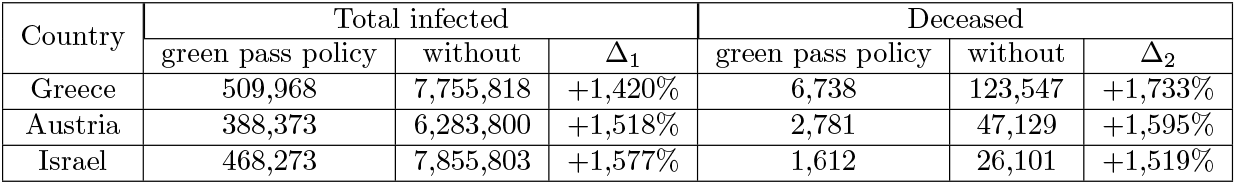
The comparison of with and without green pass policy in Greece, Austria, and Israel.

The above analyses show that the green pass policy indeed plays a crucial role in combating COVID-19 resurgence. Even the green pass policy cannot eliminate the COVID-19 spred, it can make a reasonable balance between personal freedom and national economy. Besides, a temporary strict NPIs for the vaccinated maybe also is necessary when the daily cases increase abruptly in a short time as the three countries performed a strict NPIs for the green pass holders, just like the situation of the three countries.

### Ablation study

To further evaluate the capacity of our method SEIQRD^2^, we conduct three ablation studies in Greece, Austria, and Israel. The first ablation study generates a variant SPEIQRD from the SEIQRD^2^ without the factor of green pass policy, which treats the behavior patterns of both groups as homogeneous. The second one derives a new model SVEIQRD by discarding the interactions between the unvaccinated and the vaccinated groups in SEIQRD^2^. The third ablation produces the derived model 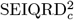 from SEIQRD^2^ by assuming the stationary efficacy of vaccination. In summary, there are three variants SPEIQRD, SVEIQRD, and 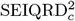, in addition to the full model SEIQRD^2^ which involves all the above aspects. The detailed information about the variants of the model is displayed in the supporting information. Besides, the results of ablation study are from independent parameters. The corresponding parameter values are from the optimization algorithm (the least-square method).

The experimental results for Greece are presented in Fig 5. Compared with the three variants SPEIQRD, SVEIQRD, and 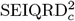, the full model SEIQRD^2^ makes more accurate forecasting of quarantined cases while its results for death cases are close to the actual values.

**Fig 5.**
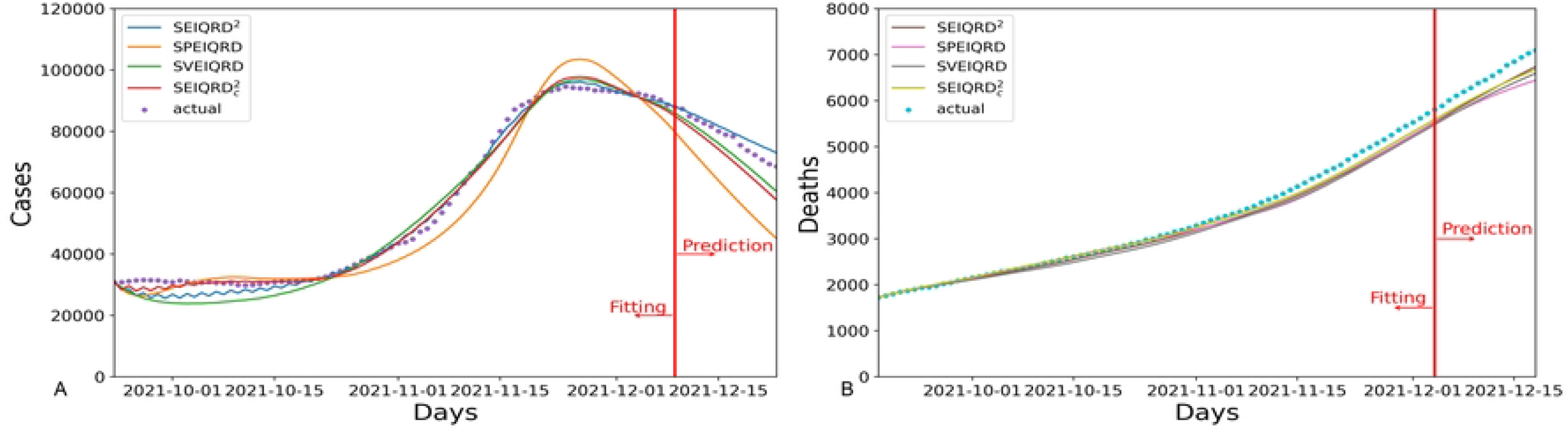
The results of four variants of our method SEIQRD^2^ in Greece. In the four variants of method, SPEIQRD ignores the impact of enforcing vaccination certification. SVEIQRD overlooks the interactions between unvaccinated and vaccinated groups. 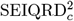 neglects the waning effectiveness of vaccine.

The results for Austria are shown in Fig 6. The three variants SPEIQRD, SVEIQRD, and 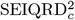 achieve the MAPE of 20.896%, 17.445%, and 24.326% respectively, while SEIQRD^2^ achieves a MAPE of 5.411% in forecasting the quarantined cases of the next 14 days. SEIQRD^2^’s evident advantage over the three variants demonstrates the significant role of NPI, vaccination, and the interaction between unvaccinated and vaccinated groups in mitigating the spread of COVID-19.

**Fig 6.**
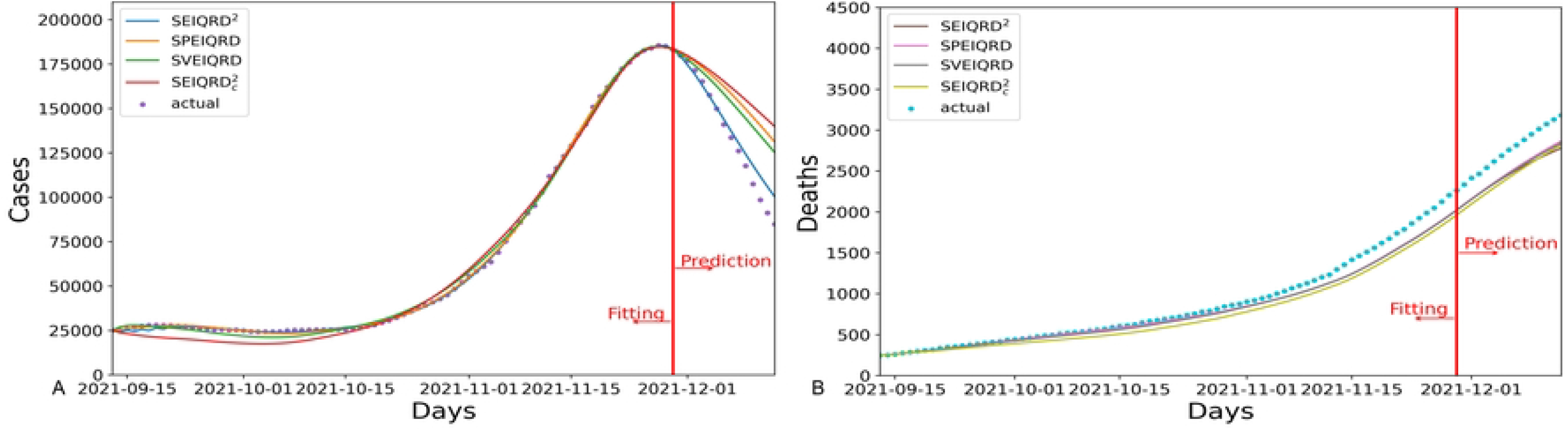
The results of four variants of our method SEIQRD^2^ in Austria.

Fig 7 displays the results for Israel. The results of SPEIQRD significantly deviate from the realistic situation, which further demonstrates that the green pass policy plays a leading role in containing the COVID-19 resurgence in Israel. In reality, Israel was the first country which carried out the green pass policy to restore its economy. Compared with Greece and Austria, Israel heavily relied on the green pass policy to contain the resurgence. This explains why the results of SPEIQRD are much worse. Among all four methods, SEIQRD^2^ obtains the least error in predicting the quarantined cases, whose MAPE is 1.292%. The forecasting performance in terms of MAPE of all models is shown in Table 6.

**Table 6.** The MAPE performance of the next 14-day prediction of cases in Greece, Austria, and Israel for the ablation study.

| Country | SEIQRD<sup>2</sup> |  | SPEIQRD |  | SVEIQRD |  | SEIQRD<sub>c</sub><sup>2</sup> |  |
| --- | --- | --- | --- | --- | --- | --- | --- | --- |
|  | quarantined | deceased | quarantined | deceased | quarantined | deceased | quarantined | deceased |
| Greece | **3.229%** | 4.853% | 22.626% | 7.014% | 6.289% | 6.184% | 8.989% | **4.676%** |
| Austria | **5.411%** | 11.133% | 20.896% | **9.999%** | 17.445% | 10.492% | 24.326% | 12.107% |
| Israel | **1.292%** | 3.167% | 18.347% | 45.994% | 13.157% | **2.557%** | 1.825% | 3.208% |

**Fig 7.**
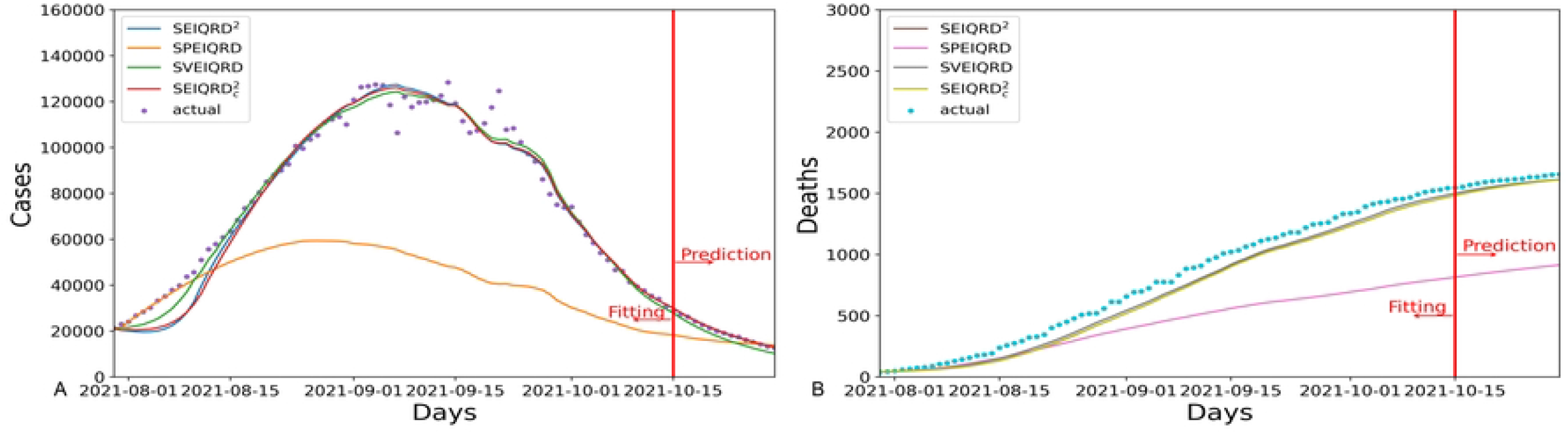
The results of four variants of our method SEIQRD^2^ in Israel.

Besides, SEIQRD^2^ achieves better results than its variant SVEIQRD, which illustrates the interaction between the two groups is a necessary factor in modeling the dynamics of COVID-19. By contrast, the poor performance of SPEIQRD emphasizes the importance of the green pass policy in the simulation of COVID-19. Although the MAPE results of both SEIQRD^2^ and SEIQRD^2^ in predicting total deaths are similar, SEIQRD^2^ makes more accurate predictions on quarantined cases than SVEIQRD. That supported the conclusion that the declined vaccine effectiveness is a crucial element in modeling of the COVID-19 pandemic.

In conclusion, these ablation studies convincingly illustrate the importance of involving the green pass policy in modeling the COVID-19. The green pass policy brought about the differences between behaviors of unvaccinated and vaccinated groups and their interactions, which complicates the COVID-19 modeling. At last, our method emphasizes the contribution of the waning efficacy of vaccine to modeling the COVID-19 infection.

## Conclusion

In this work, we develop a deterministic compartmental model SEIQRD^2^ by involving external factors. SEIQRD^2^ characterizes the resurgences caused by the coronavirus variant Delta and estimates their trends in the next period. Different from the previous SEIR-based models for COVID-19, SEIQRD^2^ incorporates three external factors, including heterogeneity of behaviors between vaccinated and unvaccinated groups, their interactions, and the waning effect of vaccine under green pass policies. We show that SEIQRD^2^ precisely matches the dynamics of COVID-19 resurgences happened in Greece, Austria, and Israel relating to the Delta variant. The modeling and forecasting results demonstrate that external factors including behavioral heterogeneity, group interaction, and vaccination status may play a crucial role in forming the dynamics of COVID-19 transmission.

In addition, the effects of these three factors are further evaluated by ablation studies. The results further elucidate that these factors improve the modeling performance in characterizing the dynamics of COVID-19 resurgence.

The study in this work further confirms that, even with substantial vaccination including boosters, NPIs may still play an essential role in suppressing or mitigating COVID-19 resurgence. At the outbreak of a new wave, quick reactions should include reasonable interventions for those vaccinated individuals in addition to other restrictions on unvaccinated people. Although the efficacy of vaccine reduces the infection risk of vaccinated people, more (frequent) activities create higher exposure for those unvaccinated to become infected, which may then become a super spreader and cause a wide spread of COVID-19. The evaluation in three countries taking different severity levels of NPIs and vaccination certification further demonstrates that it would be essential to enforce restrictions on vaccinated people during implementing vaccination certification policies.

This study is subject to several limitations. Firstly, the model parameters such as effective transmission rates, correspond to specific values, rather than a probability distributions. In the next work, we are considering the possible statistic methods for leveraging SEIR models. Secondly, our model ignores the number of hospitalized people, which may be another important external factor in deciding what time to upgrade control measures, especially for Greece and Austria co-existing with COVID-19. Once these data can be accessed by public, the related research could be made. Thirdly, the model assumes all infected individuals are detected and confirmed, which may be an ideal situation. In practice, a specific proportion of infected are asymptomatic and recovered without confirmation. In theory, the proportion is small, so it will not challenge the general results of the method. Besides, the lack of information about vaccination status also makes it hard to demonstrate the realistic change of the effective transmission rates. The problem may be solved completely when the vaccination status information is available. Even though, the values of the effective transmission rates give some insight to the role of green pass policy in COVID-19 pandemic.

Overall, this work is one of the first attempts to address the impact of green pass policy on the COVID-19 transmission. In addition, the behavior heterogeneity of unvaccinated and vaccinated groups is incorporated into the SEIR-based model to explore the possible interaction of two groups. This work could be extended to generate “what-if” scenario and help decision-makers determine appropriate NPI and mass vaccination implementations for containing future waves of infection.

## Data Availability

All data files are available from https://github.com/lzxiaohu/SEIQRD2.

## Supporting information

**S1 Dataset and code. These data and code are available at https://github.com/lzxiaohu/SEIQRD2**.

**S2 Ablation Study Models. Ablation Study Models**.

**S1 Table. The major interventions and activities reported during the resurgence caused by the Delta variant in Greece**.

**S2 Table. The major interventions and activities reported during the resurgence caused by the Delta variant in Austria**.

**S3 Table. The major interventions and activities reported during the resurgence caused by the Delta variant in Israel**.

## Acknowledgments

This work is sponsored in part by the Australian Research Council Discovery and Future Fellowship under Grant DP190101079 and Grant FT190100734..

## Author contributions

**Conceptualization:** Hu Cao

**Formal Analysis:** Hu Cao

**Funding Acquisition:** Longbing Cao

**Writing – Original Draft Preparation:** Hu Cao

**Writing – Review & Editing:** Longbing Cao

